# Designing and validating a Hereditary Spastic Paraplegia-specific Quality of Life Rating Scale (HSPQoL)

**DOI:** 10.1101/2024.07.23.24310834

**Authors:** Sue Faye Siow, Jane Fleming, Kristine Barlow-Stewart, Kishore R Kumar, Carolyn M Sue

**Affiliations:** Department of Clinical Genetics, Royal North Shore Hospital, St Leonards, NSW, Australia; Northern Clinical School, Faculty of Medicine and Health, University of Sydney, NSW, Australia; Translational Neurogenomics Group, Genomic and Inherited Disease Program, Garvan Institute of Medical Research, Darlinghurst 2010, Australia; Molecular Medicine Laboratory and Department of Neurology, Concord Hospital, Concord 2139, Australia; School of Clinical Medicine, UNSW Medicine & Health, University of New South Wales, Kensington 2052, Australia; Neuroscience Research Australia, University of New South Wales, Randwick, NSW, Australia; Department of Neurology, Prince of Wales Hospital, Randwick, NSW, Australia

**Author notes:** **Corresponding author:** Sue Faye Siow, or. **Author Contributions** All authors contributed to the study conception and design. Data collection, analysis, and writing of the first draft of the manuscript were performed by Sue-Faye Siow. All authors commented on previous versions of the manuscript. All authors read and approved the final manuscript. **Funding** Sue Faye Siow is the recipient of a National Health and Medical Research Council Postgraduate Scholarship and Australian and New Zealand Association of Neurologists Education and Research Foundation Scholarship. No other funding was received for conducting this study. **Ethics approval** This study was approved by Northern Sydney Local Health District Human Research Ethics Committee, ethics approval number 2019/ETH13187. **Data availability statement** The data that support the findings of this study are available on request from the corresponding author. The data are not publicly available due to privacy or ethical restrictions.

**Keywords:** hereditary spastic paraplegia, quality of life, patient reported outcome measure, modified Delphi, cognitive interview

## Abstract

**Background:** Patients with Hereditary Spastic Paraplegia (HSP) report reduced quality of life (QoL) compared to the general population. Generic QoL measures do not address disease-specific aspects such as spasticity, access to specialty HSP clinics, and bladder symptoms. We designed and validated an HSP-specific QoL scale (HSPQoL), intended for use in standard clinical settings and clinical trials.

**Methods:** HSP-specific items were added to the RAND 36-Item Short Form Health Survey (SF-36) to form HSPQoL. Following literature review/expert input, 23 items were presented to a panel of HSP clinicians, patients, and patient representatives (n=12) using a modified Delphi process. Items were ranked for clarity and relevance (inclusion criteria: 80% consensus). Interviews with patients (n=5) assessed suitability, comprehension, clarity, and response options to additional items. Patients completed the HSPQoL and EQ5D-5L for evaluation of construct validity (correlation, exploratory factor analysis) and test-retest reliability.

**Results:** Following the modified Delphi process, 21/23 items met the inclusion criteria. Based on cognitive interview results, items were modified (n=4), removed (n=7), or added (n=3). Sixty-one participants completed the HSPQoL and EQ5D-5L. The HSPQoL was repeated with 19 patients: 15/17 additional items moderately to strongly correlated with pre-existing SF-36 subscores (Spearman correlation 0.319-0.771, p<0.05). Exploratory factor analyses showed high percentage of variance in the first component (>45%). HSPQoL demonstrated good internal consistency (Cronbach alpha 0.94), test-retest reliability (ICC 0.957), and convergent validity with EQ5D-5L (r=0.725).

**Conclusions:** Demonstrated validity and reliability of the HSPQoL confirms consideration of its use for assessing specific QoL in individuals with HSP.

## Introduction

Hereditary Spastic Paraplegia (HSP) refers to a group of inherited neurodegenerative disorders characterized by lower limb spasticity and increased reflexes. ^1^ Studies exploring the experience of patients with HSP have identified features specific to HSP that influence overall quality of life (QoL), including spasticity, cramps, impaired mobility, pain, fatigue, bladder symptoms, poor sleep, access to treatment and support, and depression. ^2–8^ Generic health related QoL measures have been used to demonstrate reduced QoL in patients with HSP. ^2, 9, 10^ However, these measures QoL do not address HSP specific QoL aspects and may not fully represent the patient experience.

There is no curative treatment for HSP although there are several drug candidates that are ready for clinical trials. ^11^ Patient reported outcome measures (PROMs) have not been consistently used in HSP clinical trials. ^12^ Generic QoL measures, such as the 36-item short form health survey (SF-36) and the EuroQoL-5 Dimensions (EQ-5D), have previously been used in some clinical trials for patients with HSP, but only as secondary outcome measures.^13, 14^

A recent study investigating clinician and patient reported outcomes in a cohort of patients with HSP demonstrated that the generic QoL rating scale, EQ-5D was not sensitive to change over time and did not correlate with disease severity. ^9^ The SF-36 demonstrated poorer QoL in patients with HSP compared to the general population^10, 15^ and correlation with disease severity. ^15, 16^ So, while the SF-36 is a widely used, standardized and validated health related QoL measure, it has not been tested in the HSP population for sensitivity to change over time and internal consistency. As a generic measure, the SF-36 is likely to be less sensitive than an HSP-specific QoL survey as it does not capture aspects specific to HSP. There was no HSP-specific QoL scale at the time of study conception, therefore we aimed to develop the first HSP-specific QoL scale, HSPQoL. Since then, TreatHSP-QoL^17^, an HSP-specific QoL scale has been published and comparisons can therefore be made between the two scales.

## Methods

### Ethics

This study was approved by Northern Sydney Local Health District Human Research Ethics Committee, ethics approval number 2019/ETH13187. All participants provided informed written or verbal consent.

### HSPQoL design

As the SF-36 has been used in HSP patient cohorts, for our study we combined it with additional HSP-specific questions. Such an amendment to the SF-36 was used to develop the Multiple Sclerosis QoL scale (MSQoL) which has subsequently been validated in various populations. ^18, 19^ We chose to use the RAND-36 version of the SF-36 for supplementation with HSP specific items, as it is freely available, has a published scoring system, population norms, and changes are permitted. ^20^

### Literature Review

A comprehensive literature review was performed to identify common themes related to QoL in patients with HSP. The PubMed database was searched using the terms “hereditary spastic paraplegia”, “quality of life”, and “patient reported outcome measures”. Inclusion criteria were original full text research articles, published in peer reviewed journals, and written in English. Additional relevant articles from the reference lists of included articles were also reviewed. Common themes were identified from the reviewed articles. Additional items to supplement the SF36 were designed to address identified themes.

### Modified Delphi process

Panel selection: Potential panel participants were identified through the network of HSP specialists in Australia and internationally, patients of the Neurogenetics Clinic in Royal North Shore Hospital, NSW, Australia, and representatives of the HSP Research Foundation, an Australian-based HSP patient support group with members from across the world. Invitations were sent out via email and potential participants were informed of expected time for completion and number of rounds.

Round 1: The additional items were distributed to the expert panel in a two-round modified Delphi process ^21^ via email. Responses were collected using REDCap electronic data capture tools hosted at the University of Sydney. ^22^ The modified Delphi was conducted online with email invites sent to individual panel members with blinding of the identity of panel members to each other. In the first round, the items were grouped by theme and presented with the rationale for each item. Members of the panel were asked to rate the relevance and clarity of each item on a 5-point Likert scale. Free-text boxes were included to provide reasons and suggestions for modification. Participant responses were used to calculate a relevance and clarity score for each item (Supplementary Material File 2). Participant feedback in the free-text boxes was summarized.

Round 2: The scores from round 1 and summarized feedback were presented to the panel who were asked to select (1) if an item should be included, (2) if the item was not to be included, (3) if a modified version was to be included and if they agreed with the proposed modified item. The level of consensus rate for inclusion in or modification of each item was pre-determined at 80%.

### Cognitive interview process

Recruitment: Patients were recruited through the Neurogenetics Clinic, Royal North Shore Hospital, New South Wales, Australia. Eligibility criteria were a clinical diagnosis of HSP; aged older than 18 years and able to speak and read English. Eligible patients were identified through a review of the clinic patient database and contacted via phone call. Patients who participated in the modified Delphi process were not invited for the cognitive interview. Study details were emailed if patients expressed interest in participating. Patients who wanted to participate contacted the study coordinator to arrange an interview time and verbal informed consent was recorded at the beginning of the interview.

Interviews and analysis: Interviews were conducted by SFS, a specialist physician with experience consulting and counselling patients with neurological and genetic conditions. SFS was trained and supervised by a genetic counsellor and researcher with extensive experience in qualitative research methods. Structured patient interviews were conducted via phone call and audio-recorded with consent. A template for the interview transcript is available upon request. A combination of ‘think-aloud’ and ‘verbal probing’ techniques were used and an open-ended question for overall feedback accompanied each item.^23^ Interviews were transcribed verbatim for analysis. Interview responses were compiled for each item and coded into the relevant themes. ^24^ Items that had issues identified by two or more respondents were revised and additional definitions were added where required.

### Final validation step

The final HSPQoL consisting of 54 items (36 items from SF-36 and 18 additional items) was distributed via RedCap to individuals with HSP from the Neurogenetics Clinic at Royal North Shore Hospital, Sydney, Australia, and members of the HSP Research Foundation who responded to an email invitation to participate. Relevant demographic information was collected, and the EQ5D-5L^25^ scale was included to test convergent validity. Participants were then invited to repeat the HSPQoL two weeks later for test-retest reliability. RedCap responses were collated and data checking and clean up was performed. SPSS version 29,^26^ and Microsoft Excel were used for all statistical analyses. Demographics and survey scores were analyzed descriptively. The correlation of additional HSP-specific items with pre-existing SF-36 subscores was studied with Spearman correlation coefficients. Exploratory factor analysis was performed to test construct validity of SF-36 subscores with additional items. Convergent validity of HSPQoL with EQ5D-5L was tested with Pearson correlation. Cronbach alpha was used to test for internal consistency. Test-retest reliability was calculated with intraclass coefficient of scores from initial HSPQoL and repeat HSPQoL.

## Results

### Literature Review

Ten articles were reviewed, and the common themes identified. The study team designed 23 items (data available upon request) to address the five themes identified that were unique to those already assessed by the SF-36 survey: HSP specific symptoms, Visibility of HSP, Progressive nature of HSP, Access to specialized health care for HSP, Genetic nature of HSP (Supplementary Material 1- Figure 1).

**Figure 1.**
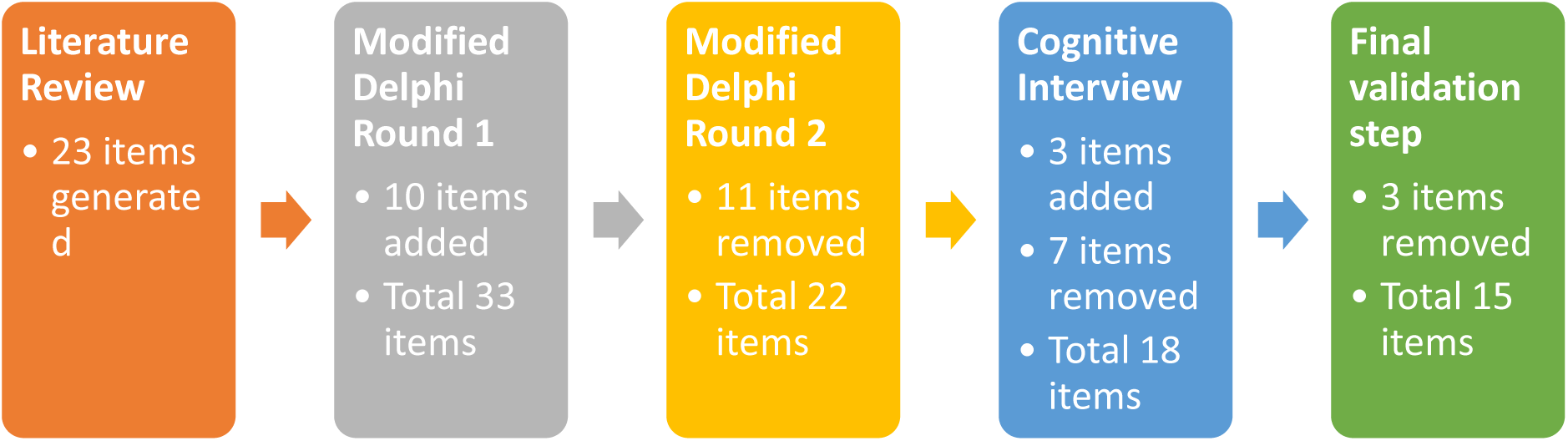
Study Design

### Modified Delphi process

Round 1: 16 email invitations were sent and the final panel consisted of 12 members (response rate 75%): HSP specialist neurologists (n=4), clinical nurse specialist (n=1), HSP patient representative (n=1), patients with HSP (n=5) and one carer for a patient with HSP. All panel members completed the modified Delphi process (Response rate 100%). The relevance and clarity scores for each question are presented in Supplementary Material 1 - Table 1. In summary, most items were ranked as moderate to high relevance (17 items with >70% relevance score), however most items were thought to be unclear (13 items with <70% clarity score). Feedback included better definition of terms used, relevance of items to respondents, relevance of respondent’s demographics, clarity of intent, and order of items.

**Table 1.** Demographics of cognitive interview participants.

| **Subject ID** | **Gender** | **Age range**<br>**(years)** | **SPRS**<br>**Score** | **Location** |
| --- | --- | --- | --- | --- |
| 1 | M | 70-79 | 22 | Metropolitan |
| 2 | M | 60-69 | 23 | Regional |
| 3 | F | 30-39 | 13 | Metropolitan |
| 4 | F | 50-59 | 21 | Regional |
| 5 | F | 70-79 | 24 | Metropolitan |

M - male, F – female, SPRS – Spastic Paraplegia Rating Scale.

Nine additional demographic questions were included to provide context for participant responses (Supplementary File 2 – List of items presented in modified Delphi round 2). Ten additional items were added based on panel feedback, and a total of 33 items were presented to the panel for Round 2 of the modified Delphi process (Figure 1).

Round 2: Response rate was 100%. 11 items that did not meet the 80% consensus rate were removed with a total of 22 items remaining (Supplementary Material 1 - Table 2). At least one item from each section met the criteria for inclusion. Feedback included review of wording, clarity of question intent, grouping together of similar questions, and standardization of response options.

**Table 2.** HSPQoL validation Participant demographics.

|  |  |  |
| --- | --- | --- |
| **Age (years)** | Mean 57.51<br>Range 28-78<br>95% CI 53.97-60.90 |  |
| **Gender n (%)** | Female 30 (49.2%) | Male 31 (50.8%) |
| **Place of residence n (%)** | Metropolitan 53 (86.9%) | Rural 8 (13.1%) |

|  | **Yes n (%)** | **No n (%)** |
| --- | --- | --- |
| **Genetic diagnosis of HSP** | 44 (72.1%) | 14 (23%) (3 don't know, 4.9%) |
| **Partner** | 42 (68.9%) | 19 (31.1%) |
| **Children** | 40 (65.6%) | 21 (34.4%) |
| **Children who have a diagnosis of HSP** | 8 | 33 |
| **Relatives with HSP** | 34 (55.7%) | 27 (44.3%) |
| **Able to walk** | 37 (60.7%) | 24 (39.3%) |
| **Access to specialised HSP clinic** | 23 (37.7%) | 38 (62.3%) |
| **Other non-HSP related health issues** | 33 (54.1%) | 28 (45.9%) |
| **Impact of non-HSP related health issues on QoL** | 20 | 13 |

### Cognitive interview

Five patients with HSP were recruited, patient demographics are listed in Table 1. Mean interview duration was 36 minutes (range 22 to 42 minutes). 22 items were presented to the interview participants for feedback. The findings of the interviews are presented in Supplementary Material 1 - Table 3. Overall, the participants reported that the additional questions covered most relevant issues. Additional comments suggested inclusion of a free-text box at the end of the survey, inclusion of information for patient support groups, inclusion of a contact person on the form as some concepts may be confusing or confronting, changes to wording of items, changes to grouping of symptoms, and modification of response options for items that may not be relevant to some respondents. Based on the results of the cognitive interview, three items were added, seven items removed, five questions were modified, and three questions were kept unchanged based on participant feedback (Figure 1). A total of 18 HSP-specific items remained for evaluation in the final validation step.

**Table 3.** SF36 subscores and additional items allocated to each subscore based on Spearman correlation and exploratory factor analysis. For specific items, please refer to HSPQoL survey available in Appendices.

| **SF36 subscore** | **Original SF36 Items** | **Additional Items** | **Themes of additional items** |
| --- | --- | --- | --- |
| **Physical Functioning** | 3-12 | 37-41 | Frequency of HSP-specific symptoms: loss of balance, falls, leg weakness, leg stiffness, spasms/cramps |
| **Social Functioning** | 20, 32 | 42-43 | Impact of HSP-specific symptoms and bladder/bowel function on QoL |
| **Pain** | 21,22 | 46 | Impact of pain on sleep |
| **Energy/Fatigue (vitality)** | 23, 27, 29, 31 | 47-49 | Impact of HSP-specific symptoms and bladder/bowel function on sleep |
| **Emotional well-being (mental health)** | 24, 25, 26, 28, 30 | 50-52 | Progressive nature, visibility, and genetic nature of HSP |
| **General Health** | 1, 33, 34, 35, 36 | 54 | Impact of access to specialized health care on QoL |
| **Role limitations due to physical health** | 13-16 | Nil |  |
| **Role limitations due to emotional problems** | 17-19 | Nil |  |
| **Health change** | 2 | Nil |  |

Of 64 individuals with HSP who responded to the email invitation to participate in the study, 61 completed the survey. Response rate was not calculated as invitations were distributed through multiple sources and the exact number of invitations received could not be determined. Invitations were then sent to 21 participants who had completed the initial HSPQoL for a re-test two weeks later with 19/21 repeat HSPQoL surveys completed (90% response rate). Participant demographics are presented in Table 2.

18 additional HSP-specific items were analyzed using Spearman correlation to map these items to the existing SF-36 subscores (Supplementary Material 1 – Table 4). All items moderately to strongly correlated with a corresponding SF36 subscores (Supplementary Material 1 - Table 4) except for items 44, 45 and 53. Those three items addressed the impact of bladder/bowel symptoms on work and lifestyle, and regarding difficulty accessing specialised healthcare for HSP. Item 54, which asked respondents to rate the impact of access to healthcare on their QoL, moderately correlated with multiple subscores but was included with the General Health subscore as theoretically this was considered the best fit.

Exploratory factor analysis was performed to determine if the additional items fit within pre-existing SF-36 subscores (Supplementary Material 1 - Table 5). All additional items had factor loadings of at least 0.40 except for items 51, which assessed how much of the time respondents wanted to hide their symptoms, and 52, which assessed their concern for passing on HSP to their children. However, these items were retained in this subscore as there was a moderate-strong correlation with the Mental Health subscore and were thought to be a good theoretical fit. Item 53, which asked if respondents had difficulty accessing health care for their HSP, could not be included in the factor analysis as it determined if respondents were able to answer item 54, which assessed how much access to health care impacted their QoL. Therefore, item 53 was moved to demographics. Items 44 and 45, which assessed the impact of bladder or bowel symptoms on work and lifestyle, did not fit with any of the SF36 subscores using Spearman correlation or exploratory factor analysis and were therefore removed. All exploratory factor analysis showed high percentage of variance in the first component (>45%) with a sharp drop off in the second component (<19.77%) demonstrating that the items best fit within their allocated subscores (Supplementary Material 1 - Table 5). Comparison of exploratory factor analysis of pre-existing SF36 subscores with and without additional items showed minimal change in percentage variance explained by first component with the addition of the HSP-specific items (Supplementary Material 1 - Table 5). This finding demonstrates that the construct of the HSPQoL remains intact and supports enrichment of the scale with the additional items. Final allocation of additional items to pre-existing SF36 subscores is presented in Table 3.

Confirmatory factor analysis was not performed as the small sample size (n=61) was insufficient for simultaneous latent modelling. Further studies with larger sample sizes will be required to determine factor loadings, which may be used to calculate summary scores, such as the Mental Health Component Score and Physical Health Component Score ^27^.

Convergent validity was determined using Pearson correlation for additional HSP-specific items and EQ5D-5L scores. EQ5D-5L scores were calculated using the Australian value set as scores are calculated based on population norms ^28^. The EQ Index scores and EQ Visual Analogue Scores were strongly correlated with total score of additional HSP items (Pearson correlation coefficient 0.725 and 0.549 respectively, p<0.001) demonstrating convergent validity.

Test-retest reliability was calculated using intraclass coefficient (ICC) scores from 19 participants who completed both the initial HSPQoL and repeat HSPQoL two weeks later. ICC was 0.957 (p<0.001), demonstrating good test-retest reliability.

The Flesch reading ease score was 70.3 and Flesch-Kincaid grade level score was 7.2 for the HSPQoL, consistent with a “fairly easy” level of readability. ^29^ This is similar to the readability scores of the SF36 survey which were 70.3 and 6.7; and equivalent to text suitable for grade 7. ^30^

In summary, 18 additional items were included in the final validation step. Three items were removed based on Spearman correlation and exploratory factor analysis results. The final HSPQoL consisted of 51 items (36 items from SF36 and 15 additional HSP-specific items).

## Discussion

In this study, we present the design and validation of the HSPQoL, an HSP-specific patient reported outcome measure intended for use in standard clinical practice and clinical trials. The HSPQoL was developed through a rigorous process of expert consensus, consumer engagement, and psychometric testing. We demonstrate content validity, construct validity, internal consistency, convergent validity, and test-retest reliability of the HSPQoL. The patient perspective was an integral component of item development with patient participation in the modified Delphi process and further item refinement with a patient cognitive interviewing approach. ^31, 32^

HSP-specific symptoms represented the majority of additional items in the HSPQoL (11/15 items). This included items addressing the frequency of lower limb spasticity, cramps, mobility, balance, bladder or bowel symptoms, and the impact of these symptoms on patients’ QoL (Table 3). By enriching a pre-existing validated health-related QoL measure with HSP-specific items, we hope to maximize the sensitivity of the HSPQoL to changes in HSP-related QoL over time. This is particularly helpful in an HSP clinical trial where the primary outcome measure should be improvement of HSP-related symptoms. The HSPQoL is a validated patient reported outcome measure that is designed to complement an HSP-specific clinician outcome measure, such as the Spastic Paraplegia Rating Scale (SPRS)^33^, in a clinical trial.

Although genetics of HSP was identified as relevant to QoL in individuals with HSP (Supplementary Material 1 - Figure 1 and Table 1), several cognitive interview participants found items regarding hereditability of HSP difficult or confronting to answer in a survey setting (Supplementary Material 1 - Table 3). This finding highlights the importance of the cognitive interview process to elicit patient feedback regarding sensitive survey items and potential impacts of those items, including potential for response bias and psychological harm. ^23^ We were able to identify the importance of discussing the genetics of HSP in a clinical setting with support from a qualified health professional. If conducted prior to an appointment, PROMs such as the HSPQoL could facilitate the discussion of sensitive topics which may not have otherwise been raised. ^34^

The progressive nature of HSP is well recognized though can be difficult to quantify, particularly in subtypes of HSP with slow progression. ^35^ We included an item to assess change in HSP symptoms, “I am able to continue enjoying leisure activities despite my HSP symptoms” in the hope of capturing subtle changes in patients’ QoL that may not be reflected on clinical rating scales, such as the SPRS, but may have an impact on their social functioning. Similarly, the visibility of HSP is another aspect that impacts patients’ QoL but is not routinely assessed in patient reported outcome measures. ^17, 36^

Despite the majority of respondents residing in a metropolitan area, most did not have access to specialized HSP services. Inclusion of an item measuring the impact of poor access to healthcare in HSP is particularly important to identify patients who will benefit from assistance linking them in with appropriate healthcare services. ^37^ In addition, measuring the impact of access to healthcare on QoL can be relevant to measures of acceptability and feasibility for interventions in clinical trials. The HSPQoL demonstrated “fairly easy” readability as measured with Flesch-Kincaid readability tests. The final survey consists of 51 items which is comparable to the MSQoL which has 54 items. ^19^ Overall, the HSPQoL is likely to accessible to most respondents or their carers.

TreatHSP-QoL is a recently published HSP-specific 25-item patient reported outcome measure^17^ developed and validated in a large HSP patient cohort (n=298). The items are grouped into five domains: General QoL and attitude to the disease, Mobility and leisure time, Medical care, Social life and occupation/work, Associated Symptoms. Although there were similar items in both the TreatHSP-QoL and HSPQoL, there were some differences. TreatHSP-QoL may be suitable for younger patient cohorts of working age and patients with complex HSP as it included items on employment, finances, and complex symptoms including speech, upper limb symptoms, and memory. On the other hand, HSPQoL may be relevant to a broader age range and varied employment statuses, and patients who are family planning as items were relevant to respondents who were working, retired, or unemployed, and addressed the genetics of HSP, but did not include items for complex HSP symptoms. As HSPQoL incorporates a pre-existing validated QoL scale, population norms for SF-36 are available for comparison with HSP patient scores and may assist stratification of HSPQoL scores. TreatHSP-QoL was developed and validated in German whilst HSPQoL was developed and validated in English. The choice between TreatHSP-QoL or HSPQoL would rely on the target participants and intended outcomes of any future HSP clinical trial.

### Limitations

There are several limitations to this study. The modified Delphi panel members and patients interviewed were native English speakers who lived in developed countries - most participants were from Australia, whilst two participants lived in Europe. Therefore, clinician and patient perspectives were limited to experiences of stakeholders from developed Western countries and healthcare systems. All patients interviewed had moderate disease severity as measured by the SPRS. Therefore, the perspective of mildly or severely affected individuals was not captured. However, the reviewed literature included perspectives of patients with varying disease severities, from different countries and language backgrounds. There may be a selection bias of participants with a particular interest in HSP research and QoL that may have influenced the results of the study.

Some participants reported that certain QoL aspects, for example concern regarding the impact of HSP on other family members and the fluctuating nature of their symptoms, were too complex to explore in a survey. We plan to include a free-text box at the end of each survey for patients to report relevant issues that are not included in the survey. This feedback will be important if HSPQoL is used as a pre-appointment PROM to inform patient management in clinic. The HSPQoL is not suitable for use in the pediatric population as it incorporates the SF-36 which is intended for use in adults^38^. In addition, the development of the additional HSP-specific questions involved only adult participants and clinicians who treat adults with HSP. Testing validity and reliability of the HSPQoL in various countries, and languages will clarify its suitability for use internationally.

## Conclusion

We have designed an HSP-specific QoL survey, the HSPQoL. Content validity was established through expert consensus and consumer engagement. Survey validity and reliability established through comprehensive psychometric testing. We intend for the HSPQoL to be used in routine clinical practice to promote discussion of patient wellbeing, and in clinical trials to measure the patient perspective of treatment outcomes.

## Supporting information

HSPQoL Survey

## Data Availability

All data produced in the present study are available upon reasonable request to the authors

## Acknowledgements

We thank the patients and their carers, clinicians and HSP Research Foundation for their participation in this study. The authors acknowledge the Statistical Consulting Service provided by Dr Stanislaus Stadlmann from the Sydney Informatics Hub, a Core Research Facility of the University of Sydney.

