## Supplementary material for "Designing and validating a Hereditary Spastic Paraplegia-specific Quality of Life Rating Scale (HSPQoL)": HSPQoL Survey

Demographic questions

| |
|---|
| What is your age? |
| How do you describe your gender identity? |
| Do you have a clinical diagnosis of Hereditary Spastic Paraplegia (HSP)? |
| Has your HSP been confirmed by a genetic test? |
| Do you have a partner? |
| Do you have children? |
| Do your children have a diagnosis of HSP? |
| Do you have relatives who have a diagnosis of HSP? |
| Are you able to walk more than 10 steps without another person's help? |
| Where do you live? |
| Do you have access to a specialised HSP clinic? |
| Do you have other health issues not related to HSP? |
| Do your other health issues not related to HSP impact your quality of life? |

HSPQoL (*items 44, 45 and 53 moved to demographic questions after final validation step; additional HSP-specific items highlighted)

| | | |
|---|---|---|
| 1 | In general, would you say your health is | 1, Excellent\|2, Very good\|3, Good\|4, Fair\|5, Poor |
| 2 | Compared to one year ago, how would you rate your health in general now? | 1, Much better now than one year ago\|2, Somewhat better now than one year ago\|3, About the same\|4, Somewhat worse now than one year ago\|5, Much worse now than one year ago |
| The following questions are about activities you might do during a typical day. Does your health now limit you in these activities? If so, how much? | | |
| 3 | Vigorous activities, such as running, lifting heavy objects, participating in strenuous sports | 1, Yes, limited a lot\|2, Yes, limited a little\|3, No, not limited at all |
| 4 | Moderate activities, such as moving a table, pushing a vacuum cleaner, bowling, or playing golf | 1, Yes, limited a lot\|2, Yes, limited a little\|3, No, not limited at all |
| 5 | Lifting or carrying groceries | |
| 6 | Climbing several flights of stairs | |
| 7 | Climbing one flight of stairs | |
| 8 | Bending, kneeling, or stooping | |
| 9 | Walking more than one kilometre | |
| 10 | Walking half a kilometre | |
| 11 | Walking 100 metres | |
| 12 | Bathing or dressing yourself | |
| During the past 4 weeks, have you had any of the following problems with your work or other regular daily activities as a result of your physical health? | | |
| 13 | Cut down the amount of time you spent on work or other activities | 1, Yes\|2, No |
| 14 | Accomplished less than you would like | 1, Yes\|2, No |
| 15 | Were limited in the kind of work or other activities | 1, Yes\|2, No |

| | | |
|---|---|---|
| 16 | Had difficulty performing the work or other activities (for example, it took extra effort) | 1, Yes\|2, No |
| | During the past 4 weeks, have you had any of the following problems with your work or other regular daily activities as a result of any emotional problems (such as feeling depressed or anxious)? | |
| 17 | Cut down the amount of time you spent on work or other activities | 1, Yes\|2, No |
| 18 | Accomplished less than you would like | 1, Yes\|2, No |
| 19 | Didn't do work or other activities as carefully as usual | 1, Yes\|2, No |
| 20 | During the past 4 weeks, to what extent has your physical health or emotional problems interfered with your normal social activities with family, friends, neighbors, or groups? | 1, Not at all\|2, Slightly\|3, Moderately\|4, Quite a bit\|5, Extremely |
| 21 | How much bodily pain have you had during the past 4 weeks? | 1, None\|2, Very mild\|3, Mild\|4, Moderate\|5, Severe\|6, Very severe |
| 22 | During the past 4 weeks, how much did pain interfere with your normal work (including both work outside the home and housework)? | 1, Not at all\|2, A little bit\|3, Moderately\|4, Quite a bit\|5, Extremely |
| These questions are about how you feel and how things have been with you during the past 4 weeks. For each question, please give the one answer that comes closest to the way you have been feeling.<br>How much of the time during the past 4 weeks... | | |
| 23 | Did you feel full of life? | 1, All of the time\|2, Most of the time\|3, A good bit of the time\|4, Some of the time\|5, A little of the time\|6, None of the time |
| 24 | Have you been a very nervous person? | 1, All of the time\|2, Most of the time\|3, A good bit of the time\|4, Some of the time\|5, A little of the time\|6, None of the time |
| 25 | Have you felt so down in the dumps that nothing could cheer you up? | 1, All of the time\|2, Most of the time\|3, A good bit of the time\|4, Some of the time\|5, A little of the time\|6, None of the time |
| 26 | Have you felt calm and peaceful? | 1, All of the time\|2, Most of the time\|3, A good bit of the time\|4, Some of the time\|5, A little of the time\|6, None of the time |
| 27 | Did you have a lot of energy? | 1, All of the time\|2, Most of the time\|3, A good bit of the time\|4, Some of the time\|5, A little of the time\|6, None of the time |
| 28 | Have you felt down? | 1, All of the time\|2, Most of the time\|3, A good bit of the time\|4, Some of the time\|5, A little of the time\|6, None of the time |
| 29 | Did you feel worn out? | 1, All of the time\|2, Most of the time\|3, A good bit of the time\|4, Some of the |

| | | |
|---|---|---|
| | | time\|5, A little of the time\|6, None of the time |
| 30 | Have you been a happy person? | 1, All of the time\|2, Most of the time\|3, A good bit of the time\|4, Some of the time\|5, A little of the time\|6, None of the time |
| 31 | Did you feel tired? | 1, All of the time\|2, Most of the time\|3, A good bit of the time\|4, Some of the time\|5, A little of the time\|6, None of the time |
| 32 | During the past 4 weeks, how much of the time has your physical health or emotional problems interfered with your social activities (like visiting with friends, relatives, etc.)? | 1, All of the time\|2, Most of the time\|3, Some of the time\|4, A little of the time\|5, None of the time |
| How TRUE or FALSE is each of the following statements for you? | | |
| 33 | I seem to get sick a lot easier than other people | 1, Definitely true\|2, Mostly true\|3, Don't know\|4, Mostly false\|5, Definitely false |
| 34 | I am as healthy as anybody I know | 1, Definitely true\|2, Mostly true\|3, Don't know\|4, Mostly false\|5, Definitely false |
| 35 | I expect my health to get worse | 1, Definitely true\|2, Mostly true\|3, Don't know\|4, Mostly false\|5, Definitely false |
| 36 | My health is excellent | 1, Definitely true\|2, Mostly true\|3, Don't know\|4, Mostly false\|5, Definitely false |
| Over the past 4 weeks, how often have you.. | | |
| 37 | Lost your balance (stumbled or tripped) | 1, Not at all\|2, Rarely (Once or twice a week)\|3, Sometimes (Most days)\|4, Often (Once or twice a day)\|5, All the time (More than twice a day)\|6, I do not walk |
| 38 | Fallen | 1, Not at all\|2, Rarely (Once or twice a week)\|3, Sometimes (Most days)\|4, Often (Once or twice a day)\|5, All the time (More than twice a day)\|6, I do not walk |
| 39 | Experienced leg weakness | 1, Not at all\|2, Rarely (Once or twice a week)\|3, Sometimes (Most days)\|4, Often (Once or twice a day)\|5, All the time (More than twice a day)\|6, I do not walk |
| 40 | Felt leg stiffness or spasticity | 1, Not at all\|2, Rarely (Once or twice a week)\|3, Sometimes (Most days)\|4, Often (Once or twice a day)\|5, All the time (More than twice a day)\|6, I do not walk |
| 41 | Felt spasms or cramps | 1, Not at all\|2, Rarely (Once or twice a week)\|3, Sometimes (Most days)\|4, Often (Once or twice a day)\|5, All the time (More than twice a day)\|6, I do not walk |

<table>
<tr><td>42</td><td>Over the past 4 weeks, how much did leg spasms, spasticity, stiffness, or cramps affect your walking or limit your activities?</td><td>1, Not at all|2, A little bit|3, Moderately|4, Quite a bit|5, Extremely|6, Not applicable</td></tr>
<tr><td>43</td><td>During the past 4 weeks, have your bladder or bowel symptoms interfered with your life (for example cannot get to the bathroom on time, have accidents)?</td><td>1, Not at all|2, A little bit|3, Moderately|4, Quite a bit|5, Extremely</td></tr>
<tr><td>44*</td><td>Have you had to stop working due to your bladder or bowel symptoms?</td><td>Yes, No</td></tr>
<tr><td>45*</td><td>Do you make lifestyle modifications (for example drink less, frequent visits to the toilet) to manage your symptoms?</td><td>Yes, No</td></tr>
<tr><td colspan="3">During the past 4 weeks, how much have the following symptoms interrupted your sleep?</td></tr>
<tr><td>46</td><td>Pain</td><td>1, Not at all|2, A little bit|3, Moderately|4, Quite a bit|5, Extremely</td></tr>
<tr><td>47</td><td>Spasms and stiffness</td><td>1, Not at all|2, A little bit|3, Moderately|4, Quite a bit|5, Extremely</td></tr>
<tr><td>48</td><td>Restless legs</td><td>1, Not at all|2, A little bit|3, Moderately|4, Quite a bit|5, Extremely</td></tr>
<tr><td>49</td><td>Bladder or bowel urgency</td><td>1, Not at all|2, A little bit|3, Moderately|4, Quite a bit|5, Extremely</td></tr>
<tr><td>50</td><td>I am able to continue enjoying leisure activities despite my HSP symptoms.</td><td>1, Definitely true|2, Mostly true|3, Don't know|4, Mostly false|5, Definitely false</td></tr>
<tr><td>51</td><td>Symptoms of HSP can be visible to others and people may act differently around you because of your symptoms.<br>Over the last 4 weeks, how much of the time have you felt that you wanted to hide your symptoms of HSP (for example avoiding situations where you have to walk or stand)?</td><td>1, All of the time|2, Most of the time|3, Some of the time|4, A little of the time|5, None of the time</td></tr>
<tr><td>52</td><td>Do you have any concern about the potential for passing on HSP to your children? (Please answer, regardless of whether or not you are planning to have children or have completed your family prior to your HSP diagnosis)</td><td>1, Not at all|2, A little bit|3, Moderately|4, Quite a bit|5, Extremely</td></tr>
<tr><td>53*</td><td>HSP is a rare condition, and it may be difficult to access health care providers with an understanding of HSP.<br>Do you have difficulty accessing health care for your HSP?</td><td>Yes, No</td></tr>
<tr><td>54</td><td>If yes, how much does this impact your quality of life?</td><td>1, Not at all|2, A little bit|3, Moderately|4, Quite a bit|5, Extremely</td></tr>
<tr><td></td><td>Any additional comments?</td><td>Free-text</td></tr>
</table>
