## Supplementary Material 2 for "Designing and validating a Hereditary Spastic Paraplegia-specific Quality of Life Rating Scale (HSPQoL)"

### Supplementary Material File 2 – Siow et al HSPQoL

**List of items presented to modified Delphi panel in Round 1.**

SECTION 1.
The aim of this section is to assess the implications of the genetic nature of HSP on quality of life.

Rationale for section: Hereditary spastic paraplegia is an inherited condition. Individuals with a genetic condition can experience concerns about passing on the condition to their children. This can affect their reproductive choices. In this section, we explore the worry, guilt and uncertainty associated with the inherited nature of HSP.

Question 1.1 explores the concerns that individuals with HSP have for their genetic relatives due to the inherited nature of the condition.
Question 1.1
How much do you worry that your siblings could have HSP?
- Not at all
- A little bit
- Moderately
- Quite a bit
- Extremely
- Not applicable (I don't have siblings)

Question 1.2 explores the worries associated with reproductive decision making for individuals with a genetic condition.

Conditional question for Question 1.2
Did/does your diagnosis of HSP influence your reproductive decisions (such as having/not having children, IVF, adoption, etc.)?
- Yes
- No
- Not applicable (I was diagnosed with HSP after I decided to/not to have children)

Question 1.2
If yes, how much worry did/do your reproductive decisions cause you?
- Not at all
- A little bit
- Moderately
- Quite a bit
- Extremely

Questions 1.3 to 1.5 address the worry and uncertainty experienced by parents associated with the inheritance risk of HSP. This risk is variable depending on which type of HSP an individual has and can be uncertain in individuals with a clinical diagnosis but no genetic diagnosis.

Conditional question for Questions 1.3 to 1.6
Do you have children?
• Yes, Please answer questions 1.3 to 1.6
• No, Please proceed to Section 2

Question 1.3
How much do you worry that your children could have HSP?
• Not at all
• A little bit
• Moderately
• Quite a bit
• Extremely

Question 1.4
If your children have HSP, how much do you worry about the impact of HSP on their future relationships?
• Not at all
• A little bit
• Moderately
• Quite a bit
• Extremely
• Not applicable (my children don't have HSP)

Question 1.5
If your children have HSP, how much do you worry about the impact of HSP on their future employment?
• Not at all
• A little bit
• Moderately
• Quite a bit
• Extremely
• Not applicable (my children don't have HSP)

Question 1.6 explores the guilt experienced by parents with HSP. Studies have identified that worries of an individual with a genetic condition passing the condition on to their children are very important to them.
Question 1.6
How much do you worry about passing on HSP to your children?
• Not at all
• A little bit
• Moderately
• Quite a bit
• Extremely
• Not applicable (I am not planning to have children)

SECTION 2.

The aim of this section is to assess the impact of symptoms specific to HSP on quality of life.

Rationale for section:
Individuals with HSP have specific symptoms that are unique to the condition. Generic QoL scales do not measure the impact of these symptoms. This section addresses areas unique to HSP identified in quality of life studies to have a substantial impact on individuals' quality of life.

Questions 2.1 to 2.6 address the most common symptoms (pain and stiffness, gait and balance, and bladder/bowel dysfunction) experienced by individuals with HSP.
Question 2.1
During the past 4 weeks, how much have you stumbled?
• Not at all
• A little bit
• Moderately
• Quite a bit
• Extremely

Question 2.2
During the past 4 weeks, how much have you fallen?
• Not at all
• A little bit
• Moderately
• Quite a bit
• Extremely

Question 2.3
During the past 4 weeks, how much of the time have you experienced leg spasms?
• All of the time
• Most of the time
• Some of the time
• A little of the time
• None of the time

Question 2.4
During the past 4 weeks, how much did leg stiffness interfere with your normal activities?
• Not at all
• A little bit
• Moderately
• Quite a bit
• Extremely

Question 2.5
During the past 4 weeks, how much of the time have your legs given way?
• All of the time
• Most of the time
• Some of the time
• A little of the time
• None of the time

Question 2.6
During the past 4 weeks, how much of the time have your bladder and/or bowel symptoms interfered with your normal social activities?
• All of the time
• Most of the time
• Some of the time
• A little of the time
• None of the time

Question 2.7 addresses the impact of HSP symptoms on sleep quality. Poor sleep quality impacts individuals' quality of life.
Question 2.7
During the past 4 weeks, how much have your symptoms (e.g. pain, spasms) interrupted your sleep?
• Not at all
• A little bit
• Moderately
• Quite a bit
• Extremely

Question 2.8 to 2.9 addresses the fluctuating nature of HSP symptoms. Individuals with HSP have reported difficulty planning their day to day activities due to the unpredictability of their condition.
Question 2.8
How much do your symptoms fluctuate?
• Not at all
• A little bit
• Moderately
• Quite a bit
• Extremely

Question 2.9
How much do fluctuations in your symptoms affect your day to day activities?
• Not at all
• A little bit
• Moderately
• Quite a bit
• Extremely
• Not applicable

SECTION 3.

The aim of this section is to assess the visibility of HSP and its impact on quality of life.

Rationale for section:
Individuals with HSP experience symptoms that can be visible to the general public, such as leg stiffness, unsteady gait and incontinence. Individuals with HSP have described difficulty explaining their symptoms in social settings. In this section, we explore the emotional impact of the visibility of HSP on individuals' quality of life.

Questions 3.1 to 3.3 explore feelings of shame and being judged based on the visibility of HSP symptoms.

Question 3.1
Over the last 4 weeks, how much of the time have you felt that you had to conceal your symptoms of HSP?
• All of the time
• Most of the time
• Some of the time
• A little of the time
• None of the time

Question 3.2
Over the last 4 weeks, how much of the time have you felt ashamed of your symptoms of HSP?
• All of the time
• Most of the time
• Some of the time
• A little of the time
• None of the time

Question 3.3
Over the last 4 weeks, how much of the time have you felt judged based on your symptoms of HSP?
• All of the time
• Most of the time
• Some of the time
• A little of the time
• None of the time

SECTION 4.

The aim of this section is the impact of the progressive nature of HSP on quality of life.

Rationale: Individuals with HSP experience progression of their symptoms that can lead to gradual restriction of their activities. This often leads to emotional distress that impacts individuals' quality of life. In this section, we explore the physical and emotional impact of the progressive nature of HSP and the associated activity limitations.

Questions 4.1 to 4.2 assess the impact of HSP on individuals' activity limitations.

Question 4.1
I am able to continue doing the things I enjoy despite my HSP symptoms.
(e.g. leisure or work activities).
• Definitely true
• Mostly true
• Don't know
• Mostly false
• Definitely false

Question 4.2
I have had to quit the activities I enjoy due to the progression of my HSP symptoms (e.g. leisure or

work activities).
• Definitely true
• Mostly true
• Don't know
• Mostly false
• Definitely false

Question 4.3 assesses feelings of fear and anxiety experienced by individuals' with HSP due to the progressive nature of the condition and uncertainty about their future.
Question 4.3
I fear for my future due to the progression of my HSP symptoms.
• Definitely true
• Mostly true
• Don't know
• Mostly false
• Definitely false

SECTION 5.

The aim of this section is to assess individuals' access to adequate healthcare and the impact of this on their quality of life.

Rationale: It is a common experience amongst individuals with HSP that there is a lack of awareness and expertise amongst healthcare providers. In this section, we explore the degree of support individuals' with HSP have from their health professionals.

Question 5.1
Do you feel you have adequate support from health professionals for the management of your HSP?
• Strongly Agree
• Mostly agree
• Neutral
• Mostly disagree
• Strongly Disagree

Question 5.2
How much does difficulty accessing health care for your HSP impact on your quality of life?
• Not at all
• A little bit
• Moderately
• Quite a bit
• Extremely

**Score calculation methods for modified Delphi Round 1**

Participants were asked:

Do you think this question is relevant to the aim of this section?

- Strongly Agree
- Mostly agree
- Neutral
- Mostly disagree
- Strongly Disagree

Do you think this question is clear?

- Strongly Agree
- Mostly agree
- Neutral
- Mostly disagree
- Strongly Disagree

Participant responses were assigned points according to the table below.

| **Participant response** | **Points** |
|---|---|
| Strongly Agree | +2 |
| Mostly Agree | +1 |
| Neutral | 0 |
| Mostly disagree | -1 |
| Strongly Disagree | -2 |

Points were added for each item and divided by maximum number of points (2*number of respondents) multiplied by 100 to produce a % score.

e.g. 4 participants “strongly agree”, 2 participants “mostly agree”, 1 participant “neutral”, 3 participants “mostly disagree”, 2 participants “strongly agree” = (4*2 + 2*1 + 1*0 + 3*-1 + 2*-2)/24*100 = 12.5%

**<u>List of items presented in modified Delphi round 2 (Highlighted items were included following this round)</u>**

Additional Demographic questions (to be included as matrix with yes/no options)

1. Do you have a clinical diagnosis of HSP?
2. Has your HSP been confirmed by a genetic test?
3. Do you have a partner?
4. Do you have children?
5. Do you have brothers or sisters to the same parents as you?
6. Do you have extended family (aunts, uncles, cousins, grandparents) who have a diagnosis of HSP?
7. Are you able to walk more than 10 steps without another person's help?
8. Where do you live? City - Y (Nearest hospital < 50km), Rural – N (Nearest Hospital >50km)
9. Do you have access to a specialised HSP clinic?

| **Original question** | **Modified question** |
|---|---|
| SECTION 2 - Symptoms specific to HSP<br>Question 2.1 to 2.3 are only for ambulant respondents<br><br>Question 2.1<br>During the past 4 weeks, how much have you stumbled?<br>• Not at all<br>• A little bit<br>• Moderately<br>• Quite a bit<br>• Extremely<br>Relevance score: 79.2%<br>Clarity score: 12.5%<br><br>Question 2.2<br>During the past 4 weeks, how much have you fallen?<br>• Not at all<br>• A little bit<br>• Moderately<br>• Quite a bit<br>• Extremely<br>Relevance score: 79.2%<br>Clarity score: 33.3% | Q2.1 Proposed modification:<br><br>Questions 2.1 to 2.4 are combined in to one question. Participants will be able to respond to each part of the question separately.<br><br>Options (a-c) for ambulant participants only.<br><br>Over the past 4 weeks, how many times have you<br>(a) lost your balance (stumbled or tripped)?<br>(b) fallen?<br>(c) experienced leg weakness?<br>(d) felt leg spasms, stiffness, spasticity or cramps?<br><br>• Not at all<br>• 0-4 times a week<br>• >4 times a week<br>• 1-4 times a day<br>• >4 times a day |

| | |
|---|---|
| Question 2.3<br>During the past 4 weeks, how much of the time have your legs given way?<br>• All of the time<br>• Most of the time<br>• Some of the time<br>• A little of the time<br>• None of the time<br>Relevance score: 70.8%<br>Clarity score: 37.5%<br><br>Question 2.4<br>During the past 4 weeks, how much of the time have you experienced leg spasms?<br>• All of the time<br>• Most of the time<br>• Some of the time<br>• A little of the time<br>• None of the time<br>Relevance score: 83.3%<br>Clarity score: 54.2% | |
| Additional question for section 2 | Over the past year, how often have you had the following injuries from falls?<br>(a) Cuts, bruises or sprains.<br>(b) Fractures (broken bones) or dislocations.<br>(c) Major injuries requiring surgery.<br><br>• 0-10 times over the past year.<br>• 11-20 times over the past year.<br>• 21-30 times over the past year.<br>• 31-40 times over the past year<br>• >40 times over the past year |
| Question 2.5<br>During the past 4 weeks, how much did leg stiffness interfere with your normal activities?<br>• Not at all<br>• A little bit<br>• Moderately<br>• Quite a bit<br>• Extremely<br>Relevance score: 91.7%<br>Clarity score: 79.2% | Q2.5 Proposed modification:<br>During the past 4 weeks, how much did leg spasms, spasticity, stiffness or cramps affect your walking or limit your activities?<br>• Not at all<br>• A little bit<br>• Moderately<br>• Quite a bit<br>• Extremely<br>• Not applicable |
| Question 2.6<br>During the past 4 weeks, how much of the time have your bladder and/or bowel symptoms interfered with your normal social activities?<br>• All of the time<br>• Most of the time<br>• Some of the time | Q2.6 Proposed modification:<br>During the past 4 weeks, have your bladder and/or bowel symptoms interfered with your life, (i.e. cannot get to the bathroom on time, have accidents, etc.) affecting<br>(a) social activities?<br>(b) ability to work? |

| | |
|---|---|
| • A little of the time<br>• None of the time<br>Relevance score: 87.5%<br>Clarity score: 58.3% | • Not at all<br>• A little bit<br>• Moderately<br>• Quite a bit<br>• Extremely |
| Question 2.7<br>During the past 4 weeks, how much have your symptoms (e.g. pain, spasms) interrupted your sleep?<br>• Not at all<br>• A little bit<br>• Moderately<br>• Quite a bit<br>• Extremely<br>Relevance score: 79.2%<br>Clarity score: 54.2% | Q2.7 Proposed modification:<br>During the past 4 weeks, how much have the following symptoms interrupted your sleep?<br>(a) pain<br>(b) spasms and stiffness<br>(c) restless legs<br>(d) bladder/bowel urgency<br><br>• Not at all<br>• A little bit<br>• Moderately<br>• Quite a bit<br>• Extremely |
| Question 2.8<br>How much do your symptoms fluctuate?<br>• Not at all<br>• A little bit<br>• Moderately<br>• Quite a bit<br>• Extremely<br>Relevance: 66.7%<br>Clarity score: 37.5% | Q2.8 Proposed modification:<br>How much do your HSP symptoms (pain, spasms, stiffness, restless legs, bladder/bowel urgency) change during the day?<br>• Not at all<br>• A little bit<br>• Moderately<br>• Quite a bit<br>• Extremely |
| Question 2.9<br>How much do fluctuations in your symptoms affect your day to day activities?<br>• Not at all<br>• A little bit<br>• Moderately<br>• Quite a bit<br>• Extremely<br>• Not applicable<br>Relevance score: 58.3%<br>Clarity score: 58.3% | Q2.9 Proposed modification:<br>Q2.9 only to appear if participants experience symptom fluctuations in Q2.8.<br>How much do changes in your HSP symptoms affect your ability to plan your day to day activities?<br>• Not at all<br>• A little bit<br>• Moderately<br>• Quite a bit<br>• Extremely |
| SECTION 3 - Progressive Nature of HSP<br>Question 3.1<br>I am able to continue doing the things I enjoy despite my HSP symptoms.<br>(e.g. leisure or work activities).<br>• Definitely true<br>• Mostly true<br>• Don't know<br>• Mostly false<br>• Definitely false<br>Relevance score: 87.5%<br>Clarity score: 79.2% | Q3.1 Proposed modification:<br>I am able to continue enjoying leisure activities despite my HSP symptoms.<br>• Definitely true<br>• Mostly true<br>• Don't know<br>• Mostly False<br>• Definitely false |

| | |
|---|---|
| Question 3.2<br>I have had to quit the activities I enjoy due to the progression of my HSP symptoms (e.g. leisure or work activities).<br>• Definitely true<br>• Mostly true<br>• Don't know<br>• Mostly false<br>• Definitely false<br>Relevance score: 66.7%<br>Clarity score: 54.2% | Q3.2 Proposed modification:<br>I have had to quit the type of work I enjoy due to progression of my HSP symptoms.<br>• Definitely true<br>• Mostly true<br>• Don't know<br>• Mostly False<br>• Definitely false<br>• Not applicable (I do not work) |
| Question 3.3<br>I fear for my future due to the progression of my HSP symptoms.<br>• Definitely true<br>• Mostly true<br>• Don't know<br>• Mostly false<br>• Definitely false<br>Relevance score: 79.2%<br>Clarity score: 75% | Q3.3 Proposed modification:<br>I remain hopeful for my future despite the progression of my HSP symptoms.<br>• Definitely true<br>• Mostly true<br>• Don't know<br>• Mostly False<br>• Definitely false |
| Section 4 - Visibility of HSP<br><br>Question 4.1<br>Over the last 4 weeks, how much of the time have you felt that you had to conceal your symptoms of HSP?<br>• All of the time<br>• Most of the time<br>• Some of the time<br>• A little of the time<br>• None of the time<br>Relevance score: 66.7%<br>Clarity score: 58.3% | Q4.1 Proposed modification:<br>Over the last 4 weeks, how much of the time have you felt that you wanted to hide your symptoms of HSP (e.g. avoiding situations where you have to walk or stand)?<br>• Not at all<br>• A little bit<br>• Moderately<br>• Quite a bit<br>• Extremely |
| Question 4.2<br>Over the last 4 weeks, how much of the time have you felt ashamed of your symptoms of HSP?<br>• All of the time<br>• Most of the time<br>• Some of the time<br>• A little of the time<br>• None of the time<br>Relevance score: 54.2%<br>Clarity score: 50% | Q4.2 Proposed modification:<br>Over the last 4 weeks, how much of the time have you felt embarrassed by your symptoms of HSP?<br>• All of the time<br>• Most of the time<br>• Some of the time<br>• A little of the time<br>• None of the time |
| Question 4.3<br>Over the last 4 weeks, how much of the time have you felt judged based on your symptoms of HSP?<br>• All of the time | Q4.3 Proposed modification:<br>Over the last 4 weeks, how much of the time have you felt people acted differently around you because of your symptoms of HSP?<br>• All of the time |

| | |
|---|---|
| • Most of the time<br>• Some of the time<br>• A little of the time<br>• None of the time<br>Relevance score: 75%<br>Clarity score: 75% | • Most of the time<br>• Some of the time<br>• A little of the time<br>• None of the time |
| SECTION 5 - Genetic Nature of HSP<br><br>Question 5.1<br>How much do you worry about passing on HSP to your children?<br>• Not at all<br>• A little bit<br>• Moderately<br>• Quite a bit<br>• Extremely<br>• Not applicable (I am not planning to have children)<br>Relevance score: 83.3%<br>Clarity score: 87.5% | Q5.1 Proposed modification:<br><br>Q5.1 only for participants who have children.<br><br>How much does the chance of passing on HSP to your children concern you?<br>• Not at all<br>• A little bit<br>• Moderately<br>• Quite a bit<br>• Extremely |
| Question 5.2<br>Conditional question<br>Did/does your diagnosis of HSP influence your reproductive decisions (such as having/not having children, IVF, adoption, etc.)?<br>• Yes<br>• No<br>• Not applicable (I was diagnosed with HSP after I decided to/not to have children)<br><br>If yes, how much worry did/do your reproductive decisions cause you?<br>• Not at all<br>• A little bit<br>• Moderately<br>• Quite a bit<br>• Extremely<br>Relevance score: 91.7%<br>Clarity score: 75% | Q5.2 Proposed modification:<br>Did/will your diagnosis of HSP influence your and your partner's decision about reproductive decisions (such as having/not having children, IVF, adoption, etc.)?<br>• Yes<br>• No<br><br>If yes, how much concern did/will your decision to have/not have children cause<br>(a) you?<br>(b) your partner?<br><br>• Not at all<br>• A little bit<br>• Moderately<br>• Quite a bit<br>• Extremely |
| Question 5.3<br>How much do you worry that your children could have HSP?<br>• Not at all<br>• A little bit<br>• Moderately<br>• Quite a bit<br>• Extremely<br>Relevance score: 83.3%<br>Clarity score: 79.2% | Q5.3 Proposed modification:<br><br>Combined with question 5.6. See proposed modification for Q5.6. |
| Question 5.4 | Q5.4 Proposed modification: |

| | |
|---|---|
| If your children have HSP, how much do you worry about the impact of HSP on their future relationships?<br>• Not at all<br>• A little bit<br>• Moderately<br>• Quite a bit<br>• Extremely<br>• Not applicable (my children don't have HSP)<br>Relevance score: 75%<br>Clarity score: 50% | If your children have HSP, how concerned are you about the impact of HSP on their:<br>(a) future relationships (partner/spouse, family, friends)?<br>(b) reproductive decisions (e.g. having/not having children, IVF, adoption, etc.) ?<br>• Not at all<br>• A little bit<br>• Moderately<br>• Quite a bit<br>• Extremely<br>• Not applicable |
| Question 5.5<br>If your children have HSP, how much do you worry about the impact of HSP on their future employment?<br>• Not at all<br>• A little bit<br>• Moderately<br>• Quite a bit<br>• Extremely<br>• Not applicable (my children don't have HSP)<br>Relevance score: 79.2%<br>Clarity score: 87.5% | Q5.5 Proposed modification:<br>If your children have HSP, how concerned are you about the impact of HSP on<br>(a) their opportunities for work?<br>(b) their need for early retirement or going on the disability pension?<br>• Not at all<br>• A little bit<br>• Moderately<br>• Quite a bit<br>• Extremely<br>• Not applicable |
| Question 5.6<br>How much do you worry that your siblings could have HSP?<br>• Not at all<br>• A little bit<br>• Moderately<br>• Quite a bit<br>• Extremely<br>• Not applicable (I don't have siblings)<br>Relevance score: 62.5%<br>Clarity score: 62.5% | Q5.6 Proposed modification:<br>How concerned are you that your blood relatives could have HSP?<br>(a) Children<br>(b) Brothers and sisters<br>(b) Extended family (cousins, etc.)<br><br>• Not at all<br>• A little bit<br>• Moderately<br>• Quite a bit<br>• Extremely<br>• Not applicable |
| SECTION 6 - Accessibility of Healthcare<br><br>Question 6.1<br>Do you feel you have adequate support from health professionals for the management of your HSP?<br>• Strongly Agree<br>• Mostly agree<br>• Neutral<br>• Mostly disagree<br>• Strongly Disagree<br>Relevance score: 83.3%<br>Clarity score: 83.3% | Q6.1 Proposed modification:<br>Do you feel you have enough support (treatments, counselling, information) from health professionals for management of your HSP?<br>• Strongly agree<br>• Mostly agree<br>• Neutral<br>• Mostly disagree<br>• Strongly disagree |
| Question 6.2 | Q6.2 Proposed modification: |

| | |
|---|---|
| How much does difficulty accessing health care for your HSP impact on your quality of life?<br>• Not at all<br>• A little bit<br>• Moderately<br>• Quite a bit<br>• Extremely<br>Relevance: 91.7%<br>Clarity: 87.5% | How much does difficulty getting health care to manage your HSP impact on your quality of life (day to day activities, social life, work life balance)?<br>• Not at all<br>• A little bit<br>• Moderately<br>• Quite a bit<br>• Extremely |

**<u>Interview Transcript</u>**

**Standard opening** at start of each interview:

“Thank you for coming to help us test out our survey questions. We are not collecting information about you. We are trying out questions on a few people so we can improve them.

I will read you the questions and I’d like to hear about what you’re thinking when you answer them. Everything that comes to mind, whether it seems important or not.

I’ll be asking you about how you come up with your answers and how you interpret the questions. We will record the interview with your consent and take lots of notes. If any question is unclear, hard to answer or does not make sense, please let me know. We will take our time within the 30 minutes. Do you have any questions before we start.

Practice think aloud: Try to visualize the place where you live, and think about how many windows there are in that place. As you count the windows, tell me what you are seeing and thinking about”

**Examples of questions asked:**

Was the question easy to understand?

How would you change the question to make it easier to understand?

Can you tell me in your own words what the question is asking?

Are you able to recall the time period asked? Is the time period appropriate?

Is this question appropriate to ask in a quality of life survey for HSP?

Overall, do you have any feedback for this question?

Was there anything else that you think should have been included in this questionnaire?
