## Supplementary Material 1 for "Designing and validating a Hereditary Spastic Paraplegia-specific Quality of Life Rating Scale (HSPQoL)"

Figure 1. Key Themes of Quality of Life in HSP identified from literature review[4-8, 10, 17, 18, 34, 35]

1. **HSP specific symptoms**
   - Pain
   - Spasticity, stiffness, cramps
   - Mobility
   - Fatigue
2. **Visibility of HSP and social and emotional impact**
   - Gait and Balance
   - Bladder and/or Bowel Dysfunction
   - Substantial fluctuation in symptoms
   - Sleep problems
   - Feeling ashamed and judged
   - Fear and frustration
   - Mood: Depression and anxiety

3. **Progressive nature of HSP and associated activity limitations**
   - Activity Limitations and quitting activities
4. **Access to specialized health care for HSP**
   - Access to reliable and practical information
   - Access to healthcare professionals with understanding of HSP
5. **Genetic nature of HSP**

Footnote. Five additional relevant articles were published since the literature review and were found to contain similar themes to the first ten articles[2, 3, 9, 36, 37].

Table 1 Relevance and clarity scores from Round 1 of modified Delphi. Calculation methods and list of items in Supplementary Material File 2.

| **Item** | **Relevance (%)** | **Clarity (%)** |
|---|---|---|
| **Section 1: Genetic nature of HSP** | | |
| **1.1** | 62.5 | 62.5 |
| **1.2** | 91.7 | 75 |
| **1.3** | 83.3 | 79.2 |
| **1.4** | 75 | 50 |
| **1.5** | 79.2 | 87.5 |
| **1.6** | 83.3 | 87.5 |
| **Section 2: Symptoms specific to HSP** | | |
| **2.1** | 79.2 | 12.5 |
| **2.2** | 79.2 | 33.3 |
| **2.3** | 83.3 | 54.2 |

| | | |
|---|---|---|
| **2.4** | 91. 7 | 79.2 |
| **2.5** | 70.8 | 37.5 |
| **2.6** | 87.5 | 58.3 |
| **2.7** | 79.2 | 54.2 |
| **2.8** | 66.7 | 37.5 |
| **2.9** | 58.3 | 58.3 |
| **Section 3: Visibility of HSP** | | |
| **3.1** | 66.7 | 58.3 |
| **3.2** | 54.2 | 50 |
| **3.3** | 75 | 75 |
| **Section 4: Progressive nature of HSP** | | |
| **4.1** | 87.5 | 79.2 |
| **4.2** | 66.7 | 54.2 |
| **4.3** | 79.2 | 75 |
| **Section 5: Access to specialized healthcare** | | |
| **5.1** | 83.3 | 83.3 |

| | | |
|---|---|---|
| **5.2** | 91.7 | 87.5 |

Table 2 Results from Round 2 of modified Delphi process. List of items available in Supplementary Material File 2.

| Item | Include question (% respondents) | Include with modification (% respondents) | Outcome |
|---|---|---|---|
| **Section 1: Symptoms specific to HSP** | | | |
| **1.1** | 50 | 100 | Include modified question |
| **1.2** | 75 | 91.67 | Include modified question |
| **1.3** | 66.67 | 83.33 | Combined with Q1.1 |
| **1.4** | 58.33 | 91.67 | Include modified question |
| **1.5** | 75 | 75 | Not included |
| **1.6** | 75 | 83.33 | Combined with Q1.1 |
| **Section 2: Progressive nature of HSP** | | | |
| **2.1 to 2.4 combined into one modified question** | 58.33 | 91.67 | Include modified question |
| **2.5** | 91.67 | 100 | Include modified question |
| **2.6** | 66.67 | 100 | Include modified question |

| | | | |
|---|---|---|---|
| **2.7** | 83.33 | 83.33 | Include modified question |
| **2.8** | 50 | 66.67 | Not included |
| **2.9** | 50 | 75 | Not included |
| **Section 3: Visibility of HSP** | | | |
| **3.1** | 58.33 | 91.67 | Include modified question |
| **3.2** | 58.33 | 75 | Not included |
| **3.3** | 75 | 66.67 | Not included |
| **Section 4: Genetic nature of HSP** | | | |
| **4.1** | 75 | 91.67 | Include modified question |
| **4.2** | 58.33 | 75 | Not included |
| **4.3** | 66.67 | 83.33 | Not included* |
| **Section 5: Access to specialized healthcare for HSP** | | | |
| **5.1** | 75 | 58.33 | Not included |
| **5.2** | 91.67 | 83.33 | Include original question |

Footnote: *Item 4.3 was not included as 5/12 respondents commented that it was vague and responses would be heavily influenced by whether it was phrased positively or negatively. The original question was phrased positively and the modified question was phrased negatively, therefore the authors felt that the nature of the question will influence an individual's response whichever way it was phrased.

Table 3 Cognitive interview results

| **Original Item** | **Reason for revision** | **Revised Item** |
|---|---|---|
| Question 1<br><br>Over the past 4 weeks, how many times have you<br>(a) Lost your balance (stumbled or tripped)?<br>(b) Fallen?<br>(c) Experienced leg weakness?<br>(d) Felt leg spasms, stiffness, spasticity or cramps?<br><br>• Not at all<br>• 0-4 times a week<br>• >4 times a weeks | [Question: 2 respondents did not realise the question was referring to "the past 4 weeks" but were able to respond once prompted.<br>[d] and [e]: 4 respondents found they had a different response for "stiffness and spasticity" symptoms to "spasms and cramps". Suggested separating the symptoms.<br>Likert scale: 3 respondents found the response options difficult to choose from, they found the prescribed frequencies too specific and 1 respondent found their symptoms varied and another found their symptoms difficult to quantify.<br>2 respondents found "non-ambulant" was difficult to understand. | Modified question 1<br><br>Over the past 4 weeks, how **often** have you<br>(a) Lost your balance (stumbled or tripped)?<br>(b) Fallen?<br>(c) Experienced leg weakness?<br>**(d) Felt leg stiffness or spasticity?**<br>**(e) Felt spasms or cramps?**<br><br>• Not at all<br>• **Rarely (Once or twice a week)**<br>• **Sometimes (Most days)**<br>• **Often (Once or twice a day)** |

| | | |
|---|---|---|
| • 1-4 times a day<br>• >4 times a day<br>• I am non-ambulant | | • **All the time (More than twice a day)**<br>**I do not walk** |
| <u>Question 2</u><br>Over the past 4 weeks, how much did leg spasms, spasticity, stiffness or cramps affect your walking or limit your activities?<br>• Not at all<br>• A little bit<br>• Moderately<br>• Quite a bit<br>• Extremely<br>• Not applicable | 1 respondent felt that "affect your walking" and "limit your activities" were different concepts<br>1 respondent felt the question was similar to question 1<br>2 respondents mentioned that their symptoms varied but felt that they were able to choose a suitable answer. | Include as is |

| | | |
|---|---|---|
| <u>Question 3</u><br>During the past 4 weeks, have your bladder and/or bowel symptoms interfered with your life, (i.e. cannot get to the bathroom on time, have accidents, etc.) affecting your<br>(a) Social activities<br>(b) Ability to work<br>• Not at all<br>• A little bit<br>• Moderately<br>• Quite a bit<br>• Extremely | [b]: 3 respondents did not work and felt that the second half of the question was not applicable to them.<br>[c]: 2 respondents have made significant modifications to their lifestyle to manage their symptoms and felt the question did not capture these changes.<br>1 respondent said that their social activities were limited by HSP symptoms other than bladder/bowel and therefore was unable to differentiate the various reasons for lifestyle limitations. | <u>Modified question 3</u><br>During the past 4 weeks,<br>(a) have your bladder and/or bowel symptoms interfered with your life (e.g. cannot get to the bathroom on time, have accidents)<br>• Not at all<br>• A little bit<br>• Moderately<br>• Quite a bit<br>• Extremely<br>**Additional questions** |

| | | |
|---|---|---|
| | | **(b) Have you had to stop working due to your bladder and/or bowel symptoms?**<br>• **Yes**<br>• **No**<br>**(c) Do you make lifestyle modifications (e.g. drink less, frequent visits to the toilet) to manage your symptoms?**<br>• **Yes**<br>• **No** |
| <u>Question 4</u><br>During the past 4 weeks, how much have the following symptoms interrupted your sleep? | 1 respondent was unsure of the meaning of “restless legs”<br>1 respondent said they did not have any symptoms as they were on baclofen | Include as is |

| | | |
|---|---|---|
| (a) Pain<br>(b) Spasms and stiffness<br>(c) Restless legs<br>(d) Bladder/bowel urgency<br>• Not at all<br>• A little bit<br>• Moderately<br>• Quite a bit<br>• Extremely | 4 respondents felt it was an easy question to answer and appropriate to leave as is. | |
| <u>Question 5</u><br>I am able to continue enjoying leisure activities despite my HSP symptoms.<br>• Definitely true<br>• Mostly true<br>• Don’t know<br>• Mostly false | 1 respondent described variability in symptoms depending on weather, and physical and mental limitations associated with HSP but was able to answer the question appropriately.<br>All respondents found the question easy to understand and answer. | Include as is |

| | | |
|---|---|---|
| • Definitely false | | |
| <u>Question 6</u><br><br>Over the last 4 weeks, how much of the time have you felt that you wanted to hide your symptoms of HSP (e.g. avoiding situations where you have to walk or stand)?<br><br>• Not at all<br>• A little bit<br>• Moderately<br>• Quite a bit<br>• Extremely<br>• Not applicable | 1 respondent thought the question was unclear, “what else would you be doing if you’re not walking or standing?”. Not something she thought of before, suggested having more explanation on the aim of the question, e.g. changed what you do to avoid being judged. Felt it was OK to include but should be better worded, perhaps with the word “self-conscious”<br><br>1 respondent needed the question repeated and took some time to understand the purpose of the question. | <u>Modified question 6</u><br><br>**Symptoms of HSP can be visible to others and people may act differently around you because of your symptoms.**<br><br>Over the last 4 weeks, how much of the time have you felt that you wanted to hide your symptoms of HSP (e.g. avoiding situations where you have to walk or stand)?<br><br>• Not at all<br>• A little bit<br>• Moderately<br>• Quite a bit<br><br>Extremely |

| | | |
|---|---|---|
| <u>Question 7</u><br><br>Did/will your diagnosis of HSP influence your and your partner's reproductive decisions (such as having or not having children, IVF, adoption, etc.)?<br><br>• Yes<br>• No<br><br>If yes, how much concern did/will your decision to have/not have children cause<br><br>(a) You?<br>(b) Your partner?<br><br>• Not at all<br>• A little bit<br>• Moderately<br>• Quite a bit | 3 respondents felt the question was not applicable to them as they had already completed their family before receiving their HSP diagnosis.<br><br>1 respondent already knew from an early age they weren't going to have children but felt the second part of the question was not applicable as the respondent had already made their decision before meeting their partner.<br><br>1 respondent suggested clarifying the partner's carrier status (e.g. for autosomal recessive HSP).<br><br>(Referring back to the Delphi process, one of the participants very clearly articulated these complexities and proposed the above question which is more inclusive to those with and without children, as well as those who were diagnosed before and after they had children. The proposed | <u>Modified question 7</u><br><br>**How much concern does the chance of passing on HSP to your children or the reality of having passed on HSP to your children cause you?**<br><br>• Not at all<br>• A little bit<br>• Moderately<br>• Quite a bit<br>• Extremely<br><br>I do not have and am not planning to have children |

| | | |
|---|---|---|
| • Extremely | question wasn't used initially as there was a high concordance from the Delphi process for including the question in its original form but further feedback from these interviews have highlighted the issues with that question.) | |
| <u>Question 8</u><br>If your children have HSP, how concerned are you about the impact of HSP on their:<br>(a) Future relationships (partner/spouse, family, friends)?<br>(b) Reproductive decisions (e.g. having/not having children IVF, adoption, etc.)?<br>• Not at all<br>• A little bit | 3 respondents found the question unclear and difficult to answer.<br>2 respondents thought the questions was hypothetical as they did not have children or their children did not have HSP.<br>1 respondent thought the question may be too confronting for parents and that a clinician should be present when the question was being asked.<br>1 respondent found the first part of the question "future relationships" not applicable as their children were already in established relationships. | Not included as questions was unclear and some respondents answered it as a hypothetical question. There was also concern that the question could be confronting. Modified question 7 addresses a similar concept. |

| | | |
|---|---|---|
| • Moderately<br>• Quite a bit<br>• Extremely<br>• Not applicable | | |
| <u>Question 9</u><br>How concerned are you that your blood relatives could have HSP?<br>(a) Children?<br>(b) Brothers and sisters?<br>(c) Extended family (cousins, etc.)?<br>• Not at all<br>• A little bit<br>• Moderately<br>• Quite a bit<br>• Extremely | 4 respondents found the question difficult to answer. There were too many complexities with regards to multiple relatives who were affected or not affected. 2 respondents found it confrontational and 1 preferred not to answer the question as it was a sensitive topic. 1 suggested this concern should be discussed in a consultation with a clinician rather than in a survey.<br>1 respondent thought it was a hypothetical question.<br>1 respondent found it difficult to quantify their worry. | Not included as concept is too complex to capture in a survey and modified question 7 addresses the concerns regarding heredity of the condition. |

| | | |
|---|---|---|
| • Not applicable | 1 respondent found it easy to answer as it was applicable to their particular family situation. | |
| Question 10<br>How much does difficulty accessing health care for your HSP impact on your quality of life?<br>• Not at all<br>• A little bit<br>• Moderately<br>• Quite a bit<br>• Extremely | 4 out of 5 respondents interpreted the question as referring to the physical logistics of attending a doctor’s appointment. 1 respondent felt the question was unclear. | Modified question 10<br>**HSP is a rare condition, and it may be difficult to access health care providers with an understanding of HSP.**<br>Do you have difficulty accessing health care for your HSP?<br>• Yes<br>• No<br>If yes, how much does this impact your quality of life?<br>• Not at all<br>• A little bit<br>• Moderately<br>• Quite a bit |

| | | |
|---|---|---|
| | | • Extremely |

Table 4 Spearman Correlation of SF-36 Subscores with additional HSP-Specific Items (* and ** indicates statistical significance p<0.05 and p<0.01)

| SF-36 Subscore/item numbers | | 37 | 38 | 39 | 40 | 41 | 42 | 43 | 44 | 45 | 46 | 47 | 48 | 49 | 50 | 51 | 52 | 53 | 54 |
|---|---|---|---|---|---|---|---|---|---|---|---|---|---|---|---|---|---|---|---|
| Physical functioning | Correlation Coefficient | .603** | .555** | .582** | .456** | .502** | .389** | .520** | .294* | .079 | .364** | .343** | .193 | .458** | .287* | .229 | .067 | .019 | .127 |
| | Sig. (2-tailed) | <.001 | <.001 | <.001 | <.001 | <.001 | .005 | <.001 | .022 | .545 | .004 | .007 | .136 | <.001 | .025 | .076 | .608 | .886 | .495 |
| Role functioning/physical | Correlation Coefficient | .312* | .401** | .408** | .413** | .253* | .361** | .299* | .176 | .119 | .223 | .245 | .147 | .341** | .151 | -.025 | .060 | .211 | .316 |
| | Sig. (2-tailed) | .014 | .001 | .001 | <.001 | .049 | .009 | .019 | .174 | .362 | .084 | .057 | .257 | .007 | .245 | .846 | .645 | .102 | .083 |
| Role functioning/emotional | Correlation Coefficient | .285* | .267* | .276* | .402** | .337** | .546** | .412** | .189 | .280* | .270* | .217 | .186 | .401** | .304* | -.270* | .190 | .203 | .517** |
| | Sig. (2-tailed) | .026 | .038 | .032 | .001 | .008 | <.001 | <.001 | .144 | .029 | .035 | .093 | .151 | .001 | .017 | .035 | .143 | .117 | .003 |

| | | | | | | | | | | | | | | | | | | | | |
|---|---|---|---|---|---|---|---|---|---|---|---|---|---|---|---|---|---|---|---|---|
| | Energy/fatigue (vitality) | Correlation Coefficient | .294* | .331** | .397** | .501** | .389** | .485** | .388** | .268* | .144 | .368** | .441** | .371** | .364** | .444** | -.090 | .251 | .063 | .315 |
| | | Sig. (2-tailed) | .021 | .009 | .002 | <.001 | .002 | <.001 | .002 | .037 | .270 | .004 | <.001 | .003 | .004 | <.001 | .491 | .052 | .632 | .084 |
| | Emotional well-being (mental health) | Correlation Coefficient | .217 | .323* | .306* | .418** | .326* | .441** | .362** | .310* | .093 | .336** | .343** | .258* | .312* | .605** | -.396** | .319* | .276* | .359* |
| | | Sig. (2-tailed) | .093 | .011 | .017 | <.001 | .010 | .001 | .004 | .015 | .478 | .008 | .007 | .044 | .014 | <.001 | .002 | .012 | .032 | .047 |
| | Social functioning | Correlation Coefficient | .291* | .304* | .481** | .527** | .471** | .548** | .475** | .206 | .105 | .417** | .441** | .466** | .456** | .532** | -.332** | .186 | .215 | .282 |
| | | Sig. (2-tailed) | .023 | .017 | <.001 | <.001 | <.001 | <.001 | <.001 | .111 | .420 | <.001 | <.001 | <.001 | <.001 | <.001 | .009 | .151 | .096 | .125 |
| | Pain | Correlation Coefficient | .166 | .254* | .245 | .296* | .353** | .458** | .503** | .168 | .126 | .771** | .490** | .384** | .466** | .376** | -.062 | .362** | -.031 | .439* |

| | | | | | | | | | | | | | | | | | | | |
|---|---|---|---|---|---|---|---|---|---|---|---|---|---|---|---|---|---|---|---|
| | | Sig. (2-tailed) | .202 | .048 | .057 | .020 | .005 | <.001 | <.001 | .197 | .333 | <.001 | <.001 | .002 | <.001 | .003 | .635 | .004 | .813 | .014 |
| | General Health | Correlation Coefficient | .342** | .399** | .559** | .503** | .412** | .370** | .230 | .171 | .028 | .260* | .419** | .158 | .269* | .251 | .027 | .040 | .027 | .398* |
| | | Sig. (2-tailed) | .007 | .001 | <.001 | <.001 | <.001 | .008 | .075 | .187 | .830 | .043 | <.001 | .223 | .036 | .051 | .835 | .758 | .836 | .027 |

Table 5 Exploratory Factor Analysis results (* Items 51 and 52 loaded on the second component however were retained in this subscore as theoretically was the best fit and showed moderate-strong correlation)

| SF36 subscore | Original Items | Additional Items | % variance explained by first component without additional items | % variance explained by first component with additional items | % variance explained by second component with additional items | Minimum factor loading |
|---|---|---|---|---|---|---|
| **Physical Functioning** | 3-12 | 37-41 | 66.18% | 54.95% | 13.95% | 0.565 |
| **Social Functioning** | 20, 32 | 42-43 | 79.47% | 62.76% | 19.77% | 0.588 |
| **Pain** | 21,22 | 46 | 91.01% | 83.38% | 10.74% | 0.884 |
| **Energy/Fatigue (vitality)** | 23, 27, 29, 31 | 47-49 | 64.33% | 48.62% | 18.05% | 0.633 |
| **Emotional well-being (mental health)** | 24, 25, 26, 28, 30 | 50-52 | 58.82% | 45.27% | 13.41% | 0.652* |
| **General Health** | 1, 33, 34, 35, 36 | 54 | 51.87% | 54.01% | 19.23% | 0.466 |
| **Role limitations due to physical health** | 13-16 | Nil | | | | |
| **Role limitations due to emotional problems** | 17-19 | Nil | | | | |
| **Health change** | 2 | Nil | | | | |
